# Spatial variability in the risk of death from COVID-19 in 20 regions of Italy

**DOI:** 10.1101/2020.04.01.20049668

**Authors:** Kenji Mizumoto, Sushma Dahal, Gerardo Chowell

## Abstract

**Objectives:** Italy has been disproportionately affected by the COVID-19 pandemic, becoming the nation with the third highest death toll in the world as of May 10^th^, 2020. We analyzed the severity of COVID-19 pandemic across 20 Italian regions.

**Method:** We manually retrieved the daily cumulative numbers of laboratory-confirmed cases and deaths attributed to COVID-19 across 20 Italian regions. For each region, we estimated the crude case fatality ratio and time-delay adjusted case fatality ratio (aCFR). We then assessed the association between aCFR and sociodemographic, health care and transmission factors using multivariate regression analysis.

**Results:** The overall aCFR in Italy was estimated at 17.4%. Lombardia exhibited the highest aCFR (24.7%) followed by Marche (19.3%), Emilia Romagna (17.7%) and Liguria (17.6%). Our aCFR estimate was greater than 10% for 12 regions. Our aCFR estimates were statistically associated with population density and cumulative morbidity rate in a multivariate analysis.

**Conclusion:** Our aCFR estimates for overall Italy and for 7 out of 20 regions exceeded those reported for the most affected region in China. Our findings highlight the importance of social distancing to suppress incidence and reduce the death risk by preventing saturating the health care system.

## Introduction

Since the first COVID-19 cases in Wuhan, China, the virus rapidly spread throughout China, and subsequently spread across all continents of the world. As of May 10, 2020 a total of 3,917,366 confirmed COVID-19 cases including 274,361 deaths have been recorded globally with 215 countries/territories/areas reporting variable disease growth rates.^1^ Moreover, the US has reported the highest number of cases (31.8%) and the highest death toll (27.5%).^1^

The severity impact of any pandemic situation like COVID-19 largely depends on the transmission rate of the disease, the capacity of the health care system, and the spectrum of clinical severity which is tied to socio-demographic factors (age, gender) and the underlying prevalence of comorbidities in the population.^2^ A better understanding of the expected influx of severe patients to the health care system during the coronavirus pandemic in different areas of the world is key to anticipate medical resources such as ICU units and ventilators which are critically needed to save the lives of severely ill patients.^2-4^

The case fatality ratio (CFR) is one of the most important epidemiological metrics to quantify the clinical severity of emerging infectious diseases such as COVID-19. ^3, 5, 6^ So far, several studies have attempted to elucidate the CFR for different population segments and geographic regions particularly based on epidemiological data from China.^7-9^ However, there is still a scarcity of studies carefully estimating the severity of the COVID-19 pandemic in populations outside China. Accumulating epidemiological data indicates that the CFR varies by geographical location, intensity of transmission, characteristics of patients such as age, sex, and comorbidity status.^8^ For example, the time-delay adjusted CFR (aCFR) for Wuhan was estimated at 12.2% compared to 4.2% for Hubei province excluding Wuhan and 0.9% in China excluding Hubei province.^6^ While rough differences in severity of the pandemic in different countries have been highlighted,^9^ there is a need to quantify spatial variability in CFR and investigate how this variability is influenced by population factors and the characteristics of the health care system.

At the time of writing, Italy was exhibiting an alarming effect of the COVID-19 pandemic with the third highest death toll after the US and the UK^1^ but the estimates of the CFR that carefully account for the delay from onset of symptoms to death are not yet available. In this study we provide estimates of the COVID-19 CFR across 20 Italian regions by linking statistical methods with publicly available daily series of confirmed cases and deaths. We also investigated the association between aCFR and sociodemographic, health care and transmission related factors using regression analysis.

## Methods

### Study setting

Italy is located in Southern Europe and there are 20 administrative regions in the country: Lombardia, Emilia Romagna, Veneto, Piemonte, Marche, Toscana, Lazio, Campania, Liguria, Friuli V.G., Sicilia, Puglia, Umbria, Molise, Trentino-Alto Adige, Abruzzo, Valle d’Aosta, Sardegna, Calabria, Basilicata.^10^ For this study we have conducted a separate analysis for Trento and Bolzano provinces within Trentino-Alto Adige region based on data availability.

The first two confirmed cases of COVID-19 in Italy were reported on 31^st^ January 2020 and had a travel history to Wuhan, China. The third case was not confirmed until February 7 and on February 22, the cumulative case count reached 9. Subsequently, the incidence trajectory rapidly increased for about 8 weeks and then gradually started to decline^11^ with daily reported incidence below 2000 cases since 30 April 2020.^12^

### Data sources

We manually retrieved the daily cumulative numbers of laboratory confirmed COVID-19 cases and deaths stratified for 21 Italian regions from the daily released report of the Ministry of Health of Italy from March 2 to May 10, 2020.^12^

For each region, we retrieved major socio-demographic and healthcare variables to explore their influence on the estimated COVID-19 aCFR across areas. We also incorporated the total number of tests, the total number of tests per population size and two transmission-related metrics: the cumulative morbidity (cumulative cases) and the cumulative morbidity rate calculated as the cumulative cases divided by the local population size. These variables are summarized in Table S2.

### Statistical analysis

The crude CFR (cCFR) is defined as the number of cumulative deaths divided by the number of cumulative cases at a specific point in time. For the estimation of CFR in real time, we employed the delay from hospitalization to death, *h_s_*, which is assumed to be given by *h_s_* = *H*(*s*) − *H*(*s*-1) for *s*>0 where *H*(*s*) is a cumulative density function of the delay from hospitalization to death and follows a gamma distribution with mean 10.1 days and SD 5.4 days, obtained from the previously published paper.^6^ Let *π_a,ti_* be the time-delay adjusted case fatality ratio on reported day *t_i_* in area *a*, the likelihood function of the estimate *π_a_,_ti_* is

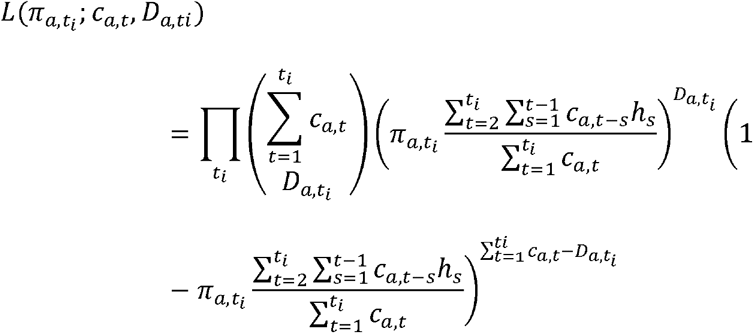

where *c_a_,_t_* represents the number of new cases with reported day t in area a, and *D_a,ti_* is the cumulative number of deaths until reported day *t_i_* in area *a*.^13, 14^ Among the cumulative cases with reported day *t* in area a, *D_a,ti_* have died and the remainder have survived the infection. The contribution of those who have died with biased death risk is shown in the middle parenthetical term and the contribution of survivors is presented in the right parenthetical term. We assume that *D_a,ti_* is the result of the binomial sampling process with probability *π_a,ti_*. We estimated model parameters using a Monte Carlo Markov Chain (MCMC) method in a Bayesian framework. Posterior distributions of the model parameters were estimated by sampling from the three Markov chains. Convergence of MCMC chains were evaluated using the potential scale reduction statistic.^15, 16^ Estimates and 95% credibility intervals for these estimates are based on the posterior probability distribution of each parameter and based on the samples drawn from the posterior distributions.

We employed multiple linear regression models to evaluate the association between regional level aCFR estimates attributable to COVID-19. A detailed description is provided in the supplement.

All statistical analyses were conducted in R version 3.6.1 (R Foundation for Statistical Computing, Vienna, Austria).

## Results

As of May 10, a total of 219,070 cases and 30,560 deaths due to COVID-19 have been reported in Italy. Moreover, the Lombardia region has reported the highest number of cases at 81,507 (32.7%) and deaths at 14,986 (49.0%) followed by Emilia Romagna with 26796 (12.2%) cases and 3845 (12.6%) deaths, and Piemonte with 28665 (13.1%) cases and 3367 (11.0%) deaths.

Figure 1 and 2 display the curves of cumulative cases and cumulative deaths in (A) Lombardia, (B) Emilia Romagna, (C) Veneto, (D) Piemonte, (E) Marche, (F) Toscana, (G) Lazio, (H) Campania, (I) Liguria, (J) Friuli V.G., (K) Sicilia, (L) Puglia, (M) Umbria, (N) Molise, (O) Trento, (P) Abruzzo, (Q) Bolzano, (R) Valle d’Aosta, (S) Sardegna, (T) Calabria, (U) Basilicata, and (V) National, over time, respectively. Cumulative cases and cumulative deaths increased rapidly in the Lombardia, Emilia Romagna, and Pimonte regions.

**Figure 1:**
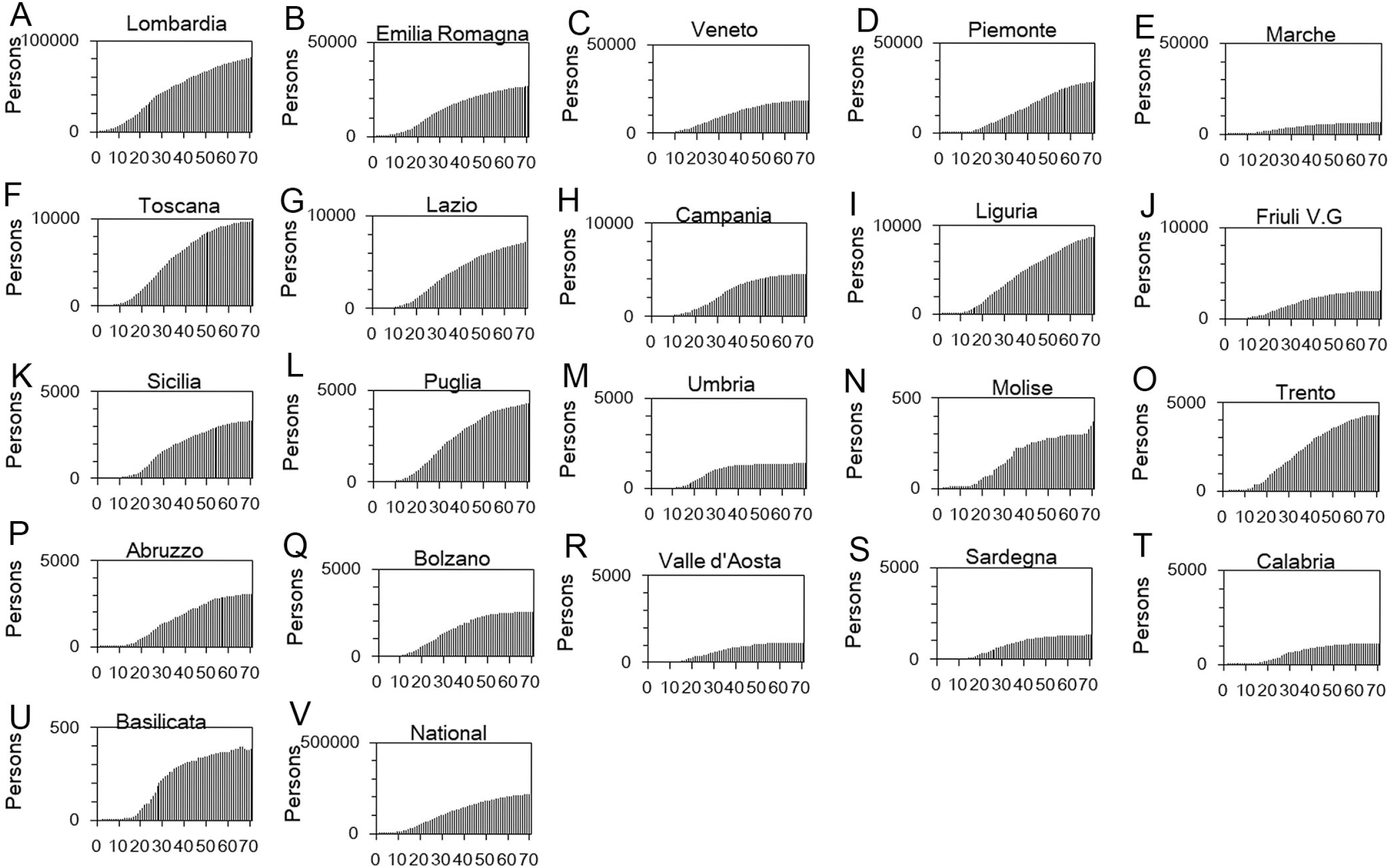
Temporal distribution of cases by region due to COVID-19, Italy, March-May 2020. Cumulative cases in (A) Lombardia, (B) Emilia Romagna, (C) Veneto, (D) Piemonte, (E) Marche, (F) Toscana, (G) Lazio, (H) Campania, (I) Liguria, (J) Friuli V.G., (K) Sicilia, (L) Puglia, (M) Umbria, (N) Molise, (O) Trento, (P) Abruzzo, (Q) Bolzano, (R) Valle d’Aosta, (S) Sardegna, (T) Calabria, (U) Basilicata, and (V) National over time. Day 1 corresponds to March 1st in 2020. As the dates of illness onset were not available, we used dates of reporting.

**Figure 2:**
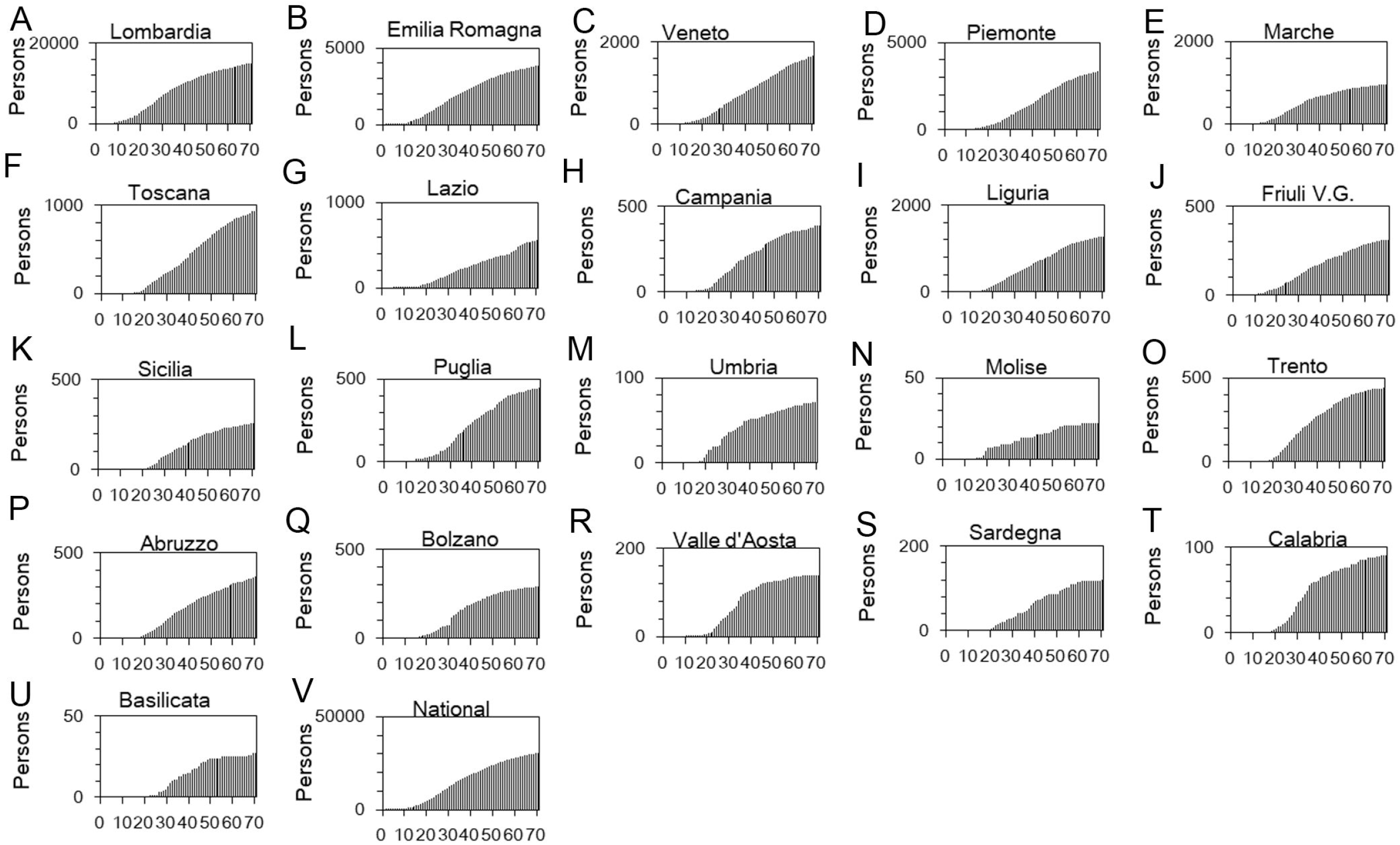
Temporal distribution of deaths by region due to COVID-19, Italy, March-May 2020. Cumulative death in (A) Lombardia, (B) Emilia Romagna, (C) Veneto, (D) Piemonte, (E) Marche, (F) Toscana, (G) Lazio, (H) Campania, (I) Liguria, (J) Friuli V.G., (K) Sicilia, (L) Puglia, (M) Umbria, (N) Molise, (O) Trento, (P) Abruzzo, (Q) Bolzano, (R) Valle d’Aosta, (S) Sardegna, (T) Calabria, (U) Basilicata, and (V) National over time. Day 1 corresponds to March 1st in 2020.

Figure 3 and 4 illustrate observed and model based posterior estimates of the cCFR and aCFR in different regions in Italy. Except for the initial days (first 5 days) our model based cCFR fitted the observed data well. For the aCFR, our model based posterior estimates are higher than the observed cCFR. Across most of the regions of Italy, the differences between cCRF and aCFR are greater in the initial 3-4 weeks and then slowly declining difference in the later stage of the epidemic. For the most affected Lombardia region, the aCFR was stable at highest point (100%) during the first 7 days (considering March 1st as day 1) and rapidly declined to 50% by day 15 and thereafter exhibited a gradual decline (about 25% by day 40). We saw a similar trend for Emilia Romagna and for the national level. For other regions such as Veneto, Marche the initial stable period was absent. For Toscana, Campania, Sicilia, Umbria, Molise, and Basilicata, the cCFR and aCFR varied slightly during the initial phase of epidemic. There was an overall downward trend of aCFR across all the regions of Italy.

**Figure 3:**
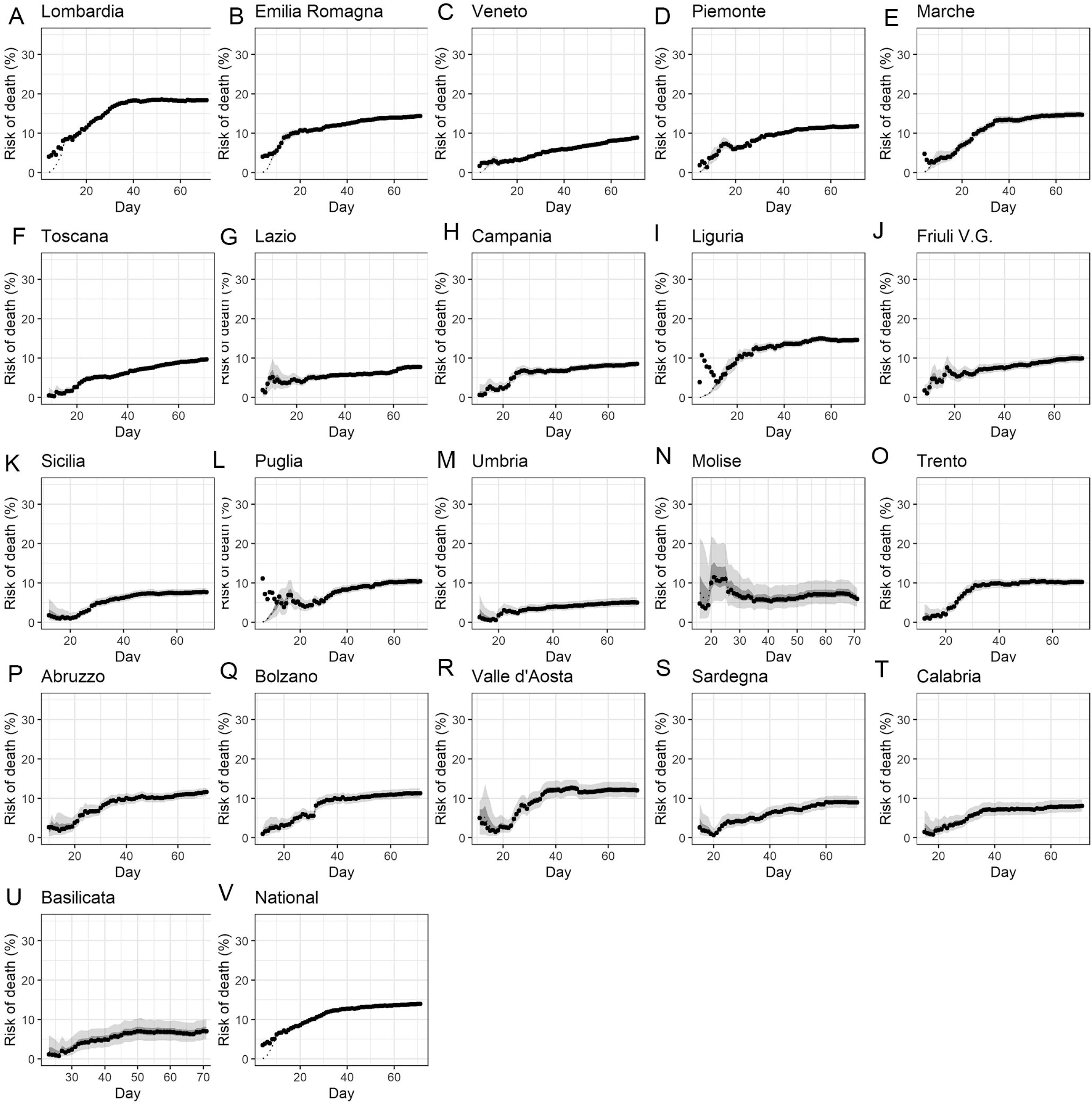
Temporal variation of risk of death caused by COVID-19 by region, Italy, March-May 2020: crude case fatality ratio (cCFR) Observed and posterior estimated of crude case fatality ratio in (A) Lombardia, (B) Emilia Romagna, (C) Veneto, (D) Piemonte, (E) Marche, (F) Toscana, (G) Lazio, (H) Campania, (I) Liguria, (J) Friuli V.G., (K) Sicilia, (L) Puglia, (M) Umbria, (N) Molise, (O) Trento, (P) Abruzzo, (Q) Bolzano, (R) Valle d’Aosta, (S) Sardegna, (T) Calabria, (U) Basilicata, and (V) National. Day 1 corresponds to March 1st in 2020. Black dots show crude case fatality ratio, and light and dark indicates 95% and 50% credible intervals for posterior estimates, respectively.

**Figure 4:**
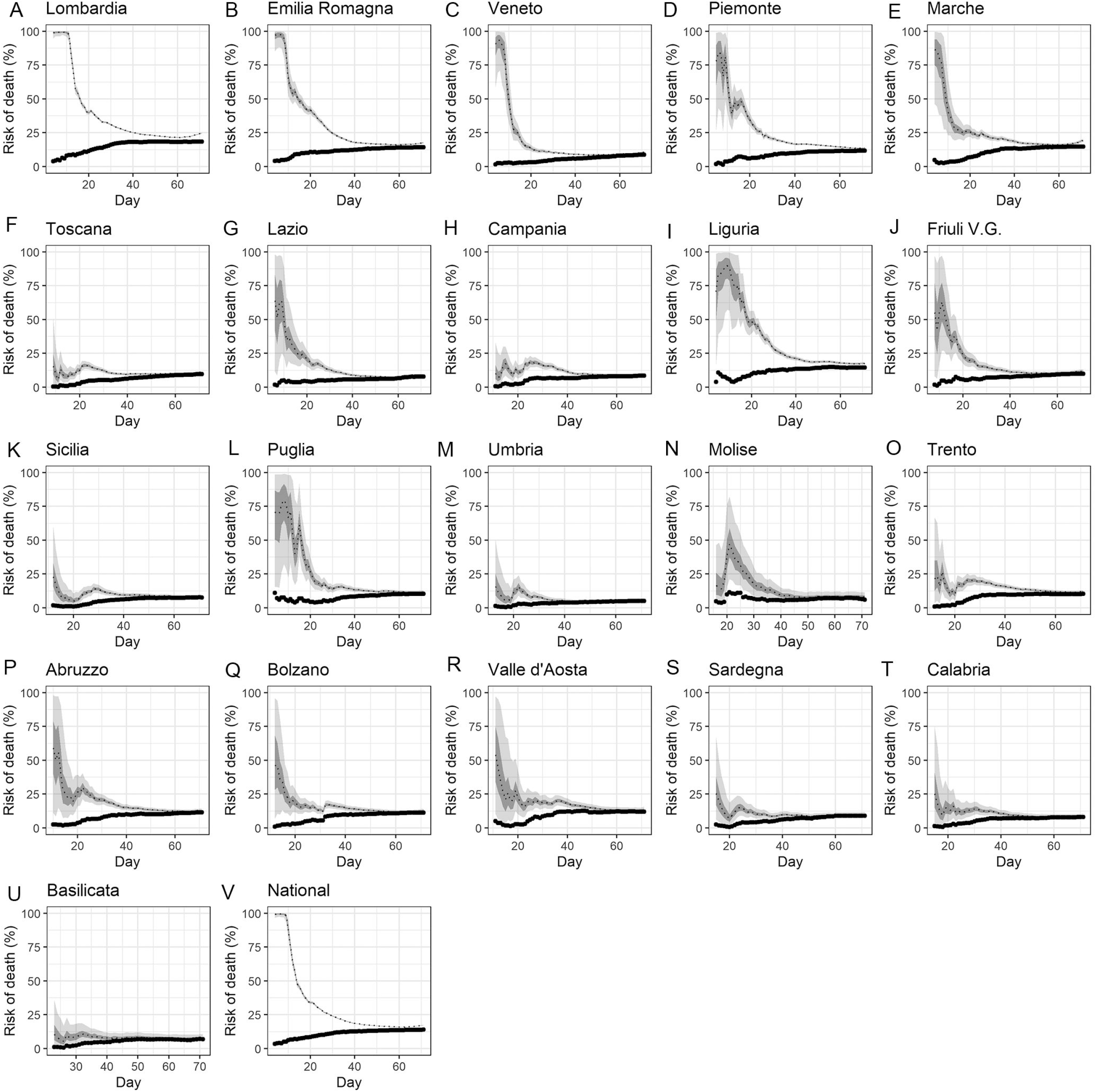
Temporal variation of risk of death caused by COVID-19 by region, Italy, March-May 2020: time-delay adjusted case fatality ratio (aCFR) Observed and posterior estimated of time-delay adjusted case fatality ratio in (A) Lombardia, (B) Emilia Romagna, (C) Veneto, (D) Piemonte, (E) Marche, (F) Toscana, (G) Lazio, (H) Campania, (I) Liguria, (J) Friuli V.G., (K) Sicilia, (L) Puglia, (M) Umbria, (N) Molise, (O) Trento, (P) Abruzzo, (Q) Bolzano, (R) Valle d’Aosta, (S) Sardegna, (T) Calabria, (U) Basilicata, and (V) National. Day 1 corresponds to March 1st in 2020. Black dots show crude case fatality ratio, and light and dark indicates 95% and 50% credible intervals for posterior estimates, respectively.

**Figure 5.**
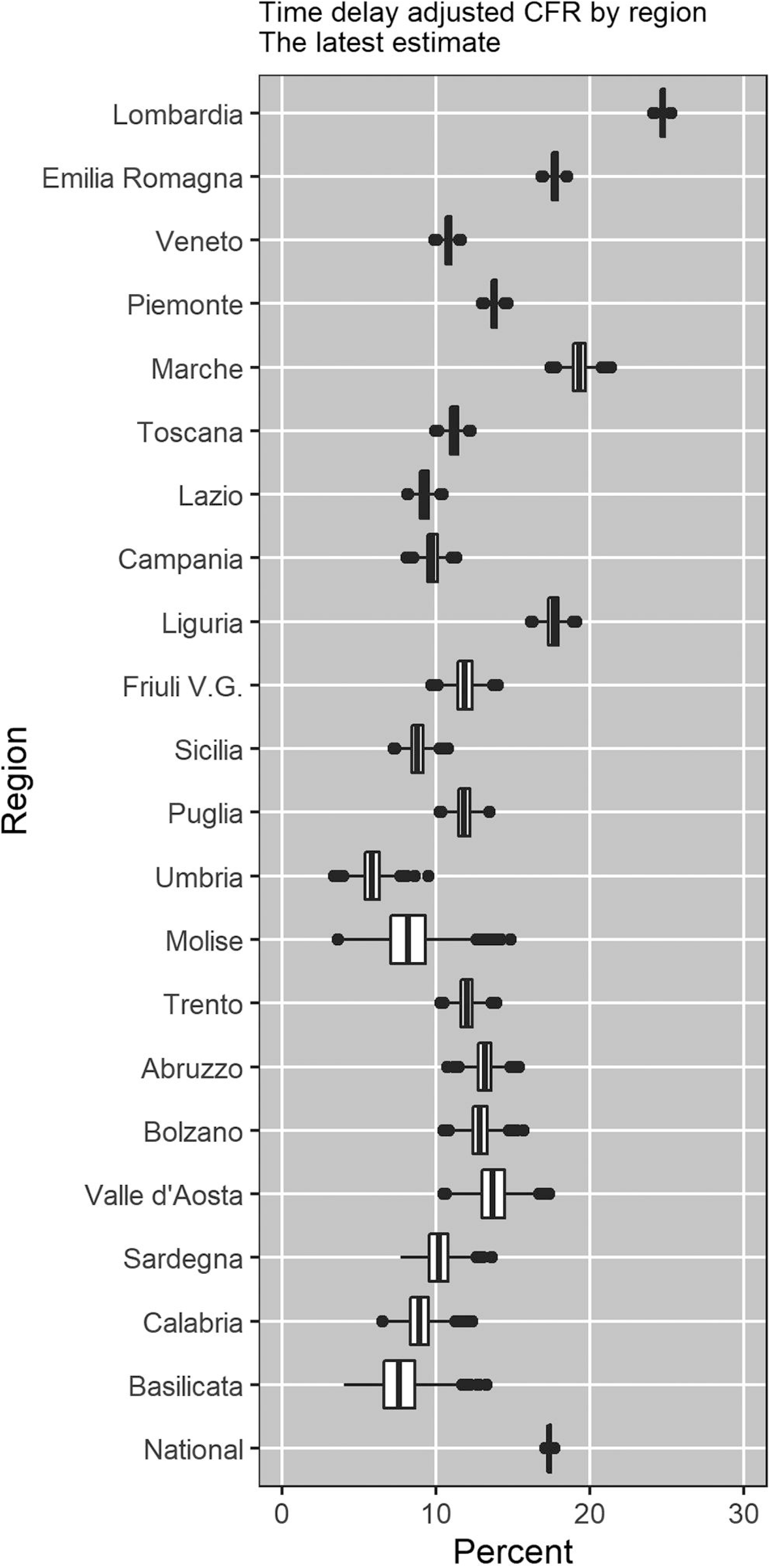
Latest estimates of time-delay adjusted risk of death caused by COVID-19 by region, 2020, Italy. Distribution of time-delay adjusted case fatality risks derived from the latest estimates (May 10, 2020) are presented. Top to bottom: Lombardia, Emilia Romagna, Veneto, Piemonte, Marche, Toscana, Lazio, Campania, Liguria, Friuli V.G., Sicilia, Puglia, Umbria, Molise, Trento, Abruzzo, Bolzano, Valle d’Aosta, Sardegna, Calabria, Basilicata and National

**Figure 6.**
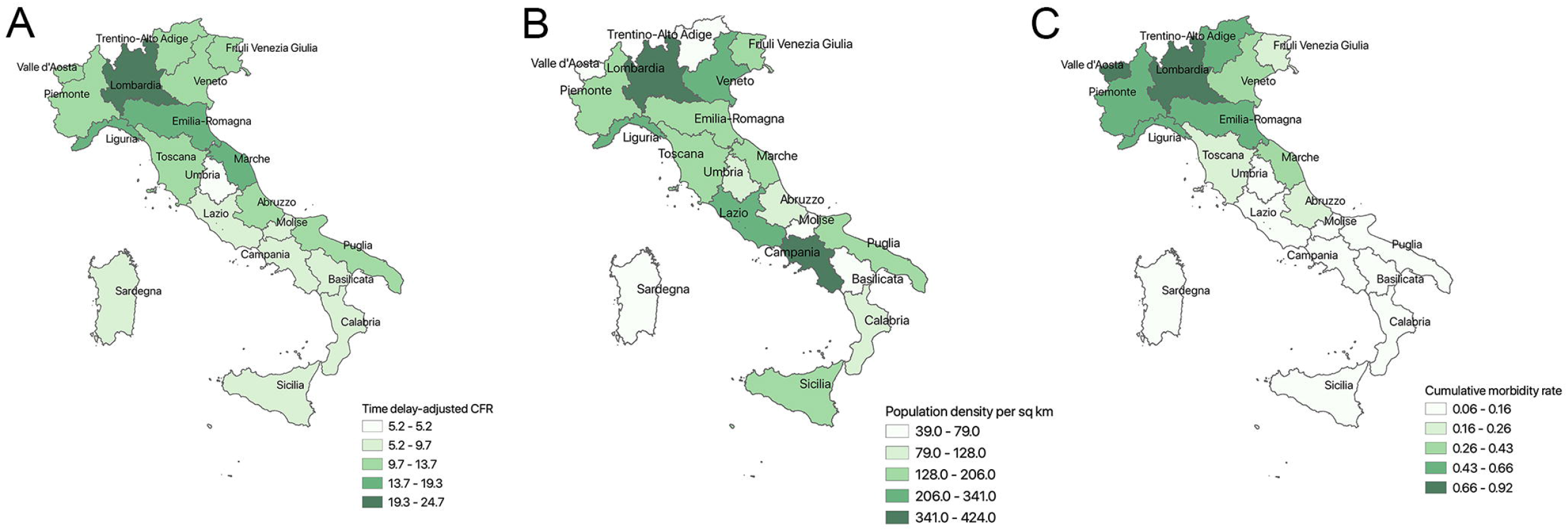
Geographical variability of COVID-19 time-delay adjusted CFR, population density, and cumulative morbidity rate across 20 regions in Italy. Distribution of time-delay adjusted case fatality risks derived from the latest estimates (May 10, 2020) are presented. Top to b (A) time –delay adjusted case fatality rate (B)Population density per square km (2019) (C) Cumulative morbidity rate in percent

A summary of the aCFR, range of median estimates and cCFR of COVID-19 across 20 regions of Italy are presented in Table 1. Lombardia had the highest aCFR of 24.7% (95% credible interval: 24.4, 25.1] followed by Marche (19.3%) [95% CrI: 18.2, 20.5], Emilia Romagna (17.7%) [95% CrI: 17.2, 18.3] and the Liguria (17.6%) [95% CrI: 16.8, 18.6] (Table 1, figure 3). The Umbria region exhibited the lowest aCFR (5.2%) [95% CrI: 4.0, 6.4] (Table 1).

**Table 1.**
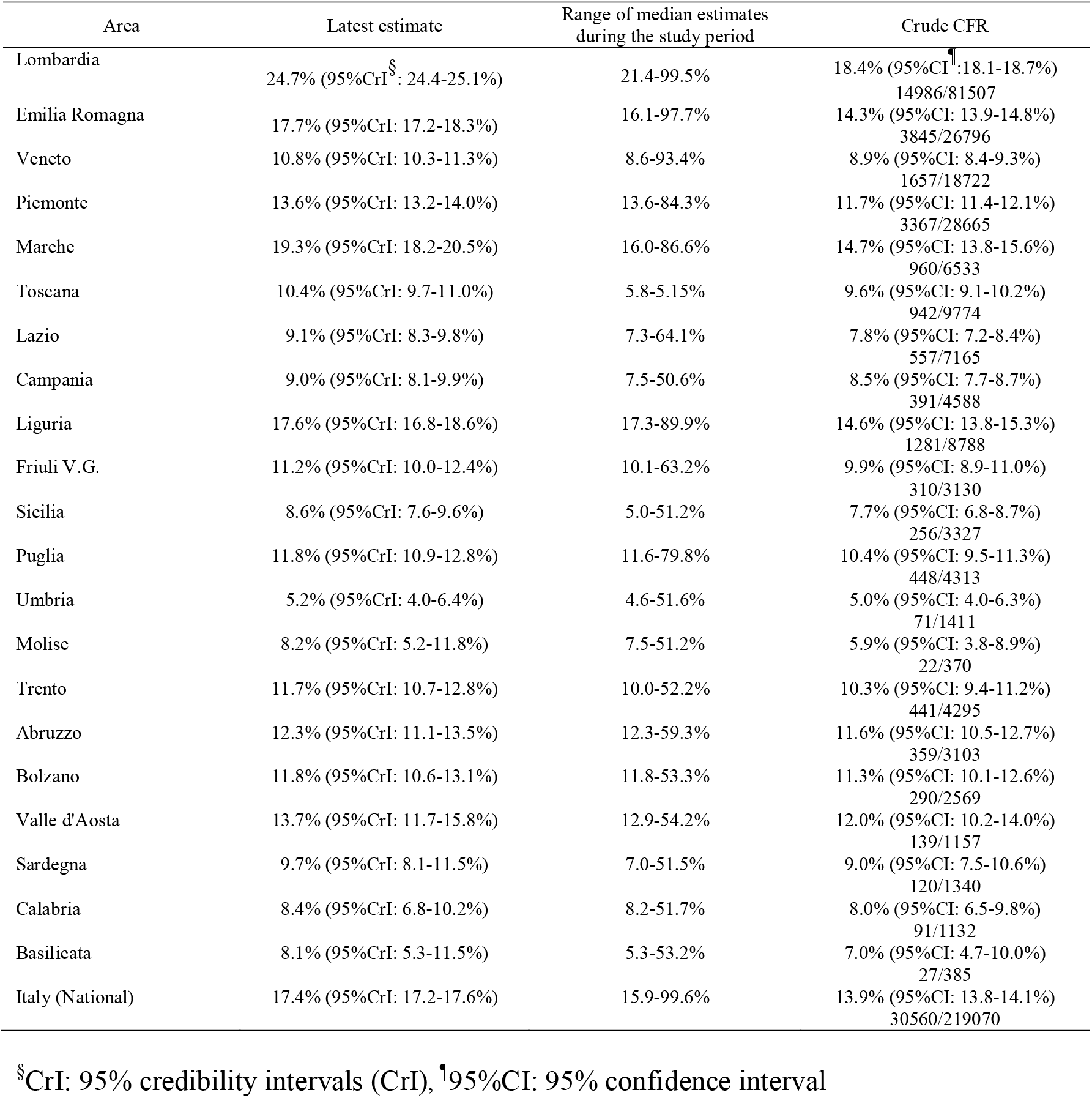
Summary results of time-delay adjusted case fatality ratio of COVID-19 by region in Italy, 2020 (As of May 10, 2020)

Regions with higher population density and those with a higher cumulative morbidity rate tended to exhibit higher aCFR estimates. These two predictors explained 58% variability (Adjusted R^2^=0.58) in the severity of pandemic across the Italian regions (P<0.05) (Table S1). Figure 7 displays the geographic distribution of the aCFR, population density per square km, and the cumulative morbidity rate across 20 Italian regions. The scatter plots and the correlation coefficients between aCFR and the variables included in the regression analysis are shown in Figure S1. Figure S2 displays the scatter plot of cumulative cases per total tests and cumulative morbidity rate for different regions of Italy. Because cumulative morbidity per total number of tests was statistically associated with cumulative morbidity rate, it was excluded due to multicollinearity (p value<0.00, r = 0.85) (Figure S2).

## Discussion

In this paper, we have estimated the time delay adjusted case fatality ratio of COVID-19 for 20 regions of Italy. Our latest estimate of aCFR in Italy varied substantially across regions with the highest value in the Lombardia region (24.7%) in the Northwest and the lowest in the Umbria region (5.2%) in the Central Italy. The aCFR estimate for the national level was 17.4%. A total of 12 administrative regions had aCFR estimates greater than 10%. Our results emphasize the need to generate real-time regional severity estimates to focus mitigation efforts and allocate medical resources that help ameliorate the burden on strained or overwhelmed health care infrastructures.

We found that the regions in Northern Italy were the most affected compared to regions in Southern Italy including Islands. The aCFR estimates across 7 administrative regions: Lombardia, Emilia Romagna, Piemonte, Marche, Liguria, Abruzzo, and Valle d’Aosta were higher than the aCFR estimates for Wuhan (12.2%) (6) and Korea (1.4%).^17^

Results of the multivariate analysis indicate that population density and cumulative morbidity rate are statistically associated with aCFR in Italy, which underscore the importance of social distancing and the need to suppress the incidence curve in order to avoid saturating the health care system and reduce the death risk. We found a statistically significant association of cumulative morbidity per total number of tests and cumulative morbidity rate. This is likely attributable to the different testing strategies implemented during the early transmission phase and in the later phase of epidemic.^18^

When we compare the aCFR of the most affected regions in Italy and China, the estimate for Italy is about twice the estimate for China (24.7% vs 12.2 %).^6^ This difference may be partly explained by the demographic structure of the two countries as suggested in a previous study,^18^ namely Italy has an older population compared to China. In 2019, approximately 23% of the Italian population was 65 years and older^18^ compared to 12.6% in China.^19^ Other factors behind the differences in the CFR estimates could be associated with the timing and intensity of public health and social measures such as ‘lockdown’ measures. In Wuhan, China aggressive lockdown measures were put in place for about 3 weeks after the report of first COVID-19 case.^20^ In Italy, Northern provinces were put under lockdowns only 5 weeks after the first recorded cases. Yet, the extent and guidelines of the lockdown were not clearly defined.^21^ Similarly, different testing strategies may have also influenced differences in CFR. Likewise, in the early phase of epidemic, there was an extensive testing strategy in Italy that included both symptomatic cases and their asymptomatic contacts but later more strict testing policies prioritized more severe suspected cases requiring hospitalization.^18^

In our study, as the epidemic progressed, we saw a downward trend in the aCFR for most of the regions in Italy. For Lombardia, Emilia Romagna and for national level, there was also an initial phase with steady high-level CFR which was relatively longer for Lombardia region compared to Emilia Romagna. A previous study on COVID-19 using data from China has also found the declining trend of aCFR for Hubei province excluding Wuhan.^6^ This trend was also reported for the 2015 MERS outbreak in South Korea in which the risk of death was significantly associated with older age and underlying health condition.^22^ In the early phase of the outbreak of an emerging infectious disease like MERS and COVID-19, the detection rate of mildly symptomatic cases is low and only patients who have serious conditions are confirmed due to hospitalization as happened in Wuhan^23,6^ and South Korea^22^. However the downward trend of CFR in the later phase of epidemic suggests both an improvement in epidemiologic surveillance and a decline in the proportion of vulnerable patients.^6^ Because of the decline in the proportion of vulnerable patients and an increased detection of mildly symptomatic cases, the epidemic might be prolonged unless strict social/physical distancing measures are applied.^6^

Our findings underscore the utility of real-time severity estimates to guide the urgent allocation medical resources in highly affected regions and the appropriate planning and supplies procurement in the other regions of Italy with a focus on medical care delivery to the most vulnerable populations with the highest risk of poorer disease outcomes due to COVID-19 such as patients categorized as critical, the elderly, and those with multiple comorbidities including cardiovascular disease, hypertension, and diabetes.^8^ Similarly, social distancing measures are critical to prevent the health care system from overloading to a breaking point. After a lockdown that lasted for about two months throughout the country, Italy started to ease movement restrictions on May 4, 2020. However, this should be conducted cautiously by putting in place the necessary infrastructure for tracing, testing, isolation and treatment in place to reduce the likelihood that the disease resurges.^24^

Our study is not exempted from limitations. The preferential ascertainment of severe cases bias in COVID-19 may have spuriously increased our estimate of CFR,^25^ which is a frequent caveat in this type of studies.^26, 27^ Similarly, for a disease like COVID-19 where transmission is characterized by a rapid growth phase in case incidence, but the infection-death time is long (ranges from 2 to 8 weeks),^8^ our CFR estimate could have been affected by delayed reporting bias.^25, 28^ Similarly, our data on number of cases reflects the date of reporting and not the date of onset of illness.

## Conclusion

The risk of death due to COVID-19 in Italy was estimated at 17.4% with varying rates across 20 regions. Our estimates of time delay adjusted CFR was as high as 24.7% in Lombardia, in Northwest Italy and as low as 5.2% in the Umbria region, located in Central Italy. Importantly, 12 out of the 20 regions exhibited aCFR values greater than 10% and the estimates for 7 regions exceeded previous estimates for the most affected regions in China and Korea. Our findings underscore the importance of social distancing to mitigate the incidence curve to reduce the risk of death from COVID-19, which we found to be significantly associated with the cumulative morbidity rates and population density.

## Data Availability

The data that support the findings of this study are available from the corresponding author, GC, upon reasonable request.

## Acknowledgments

KM acknowledges support from the Japan Society for the Promotion of Science (JSPS) KAKENHI (Grant Number 18K17368 and 20H03940) and from the Leading Initiative for Excellent Young Researchers from the Ministry of Education, Culture, Sport, Science & Technology of Japan. GC acknowledges support from NSF grant 1414374 as part of the joint NSF-NIH-USDA Ecology and Evolution of Infectious Diseases program.

## Conflict of interest

The authors declare no conflicts of interest.

### Additional files

**Additional file 1:**

**Appendix. Table S1.** Multivariate models of time-adjusted case fatality ratio as a function of major socio-demographic, and healthcare variables for 21 regions in Italy. **Table S2.** The range of major socio-demographic, and healthcare variables for 21 regions in Italy used in our analyses (linear regression). **Figure S1**. Scatter plots of time-delay adjusted CFR and other variables with correlation coefficients. **Figure S2**. Scatter plot of cumulative morbidity rate and cumulative cases per total tests with correlation coefficients.

## Notes

### Competing Interest Statement

The authors have declared no competing interest.

